# Longitudinal Measures of Cardiovascular Risk Factors Are Associated with Subfertility in the Coronary Artery Risk Development in Young Adults (CARDIA) Study

**DOI:** 10.1101/2025.01.08.25320210

**Authors:** Catherine Kim, Duke Appiah, Zhe Yin, Pamela J. Schreiner, Cora E. Lewis, Megan M. McLaughlin, Adrienne N. Dula, David S. Siscovick, Heather Huddleston

**Affiliations:** Departments of Medicine, Obstetrics & Gynecology, and Epidemiology, Ann Arbor, MI; Department of Public Health, Texas Tech University Health Sciences Center, Lubbock, TX; Department of Biostatistics, MD Anderson Cancer Center, Houston, TX; Division of Epidemiology and Community Health, University of Minnesota, Minneapolis, MN; Department of Epidemiology, University of Alabama at Birmingham, Birmingham, AL; Division of Cardiology, Department of Medicine, University of California, San Francisco, CA; Department of Neurology, Dell Medical School at The University of Texas Austin, Austin, TX; New York Academy of Medicine, New York City, NY; Department of Obstetrics & Gynecology, University of California, San Francisco, CA

**Author notes:** <u>Corresponding author</u>: Catherine Kim, MD MPH, 2800 Plymouth Road, Building 16, Room 416E, Ann Arbor, MI 48109.

## Abstract

**Introduction:** Previous reports have noted associations between subfertility in women and increased risk of cardiovascular disease (CVD) events in later life. However, reports conflict regarding the associations between subfertility and CVD risk factors. Using data from a population-based cohort of Black and White women, we examined the association between longitudinal assessments of CVD risk factors and subfertility.

**Methods:** The Coronary Artery Risk Development in Young Adults (CARDIA) study is a prospective cohort of Black and White women who have undergone repeated assessment of CVD risk factors beginning at study baseline (1985-1986). Risk factors included cigarette smoking, body mass index (BMI), blood pressure, lipid levels, glucose, and C-reactive protein. At approximately 40 years of age, an ancillary study assessed histories of subfertility. We used generalized estimating equations with a logit link model to examine associations between subfertility (dependent variable) and repeated CVD risk factors (independent variables), with adjustment for age, race, center, and education level in 1107 women.

**Results:** Cigarette use and higher levels of BMI, glucose, and triglycerides and lower levels of high-density lipoprotein cholesterol (HDL) were associated with subfertility after adjustment for age, race, and education. In multivariable models which included all of these risk factors, cigarette use (odds ratio [OR] 1.004, 95% confidence interval [CI] 1.002, 1.006, p<0.0001) and HDL (OR 0.99, 95% CI 0.99, 0.995, p=0.046) were still associated with subfertility, but associations with BMI, glucose, and triglycerides were no longer significant.

**Conclusions:** Women with subfertility histories have adverse CVD risk factors across the reproductive lifespan. Cigarette use is a strong risk factor for infertility.

## Introduction

It is well-established that a history of infertility a.k.a. subfertility in women is associated with increased risk of cardiovascular disease (CVD) [1-4]. Possible mechanisms include adverse CVD risk factor levels among women with subfertility [5]. However, previous reports conflict regarding the pattern of associations. Higher body mass index (BMI) [4-7] was associated with subfertility in some but not all [8] studies. Associations between subfertility and lipid levels was significant in some studies [4, 6] but not in others [5, 7, 8]. Findings regarding glucose [4, 5, 7, 8] and inflammatory markers [9, 10] were also contradictory. It was unclear whether associations were more pronounced in younger individuals but less pronounced for older (>35 years) women [5, 11].

The studies regarding CVD risk factor profiles among women with histories of subfertility may conflict due to measurement of CVD risk factors at different reproductive stages or different study inclusion criteria. Previous reports have typically measured CVD risk factors at a single point in time. However, the difference between CVD risk factor profiles among women with and without subfertility may differ by age [11]. In addition, multiple studies of subfertility examine select populations, including women seeking fertility services; health professionals; pregnant women; and Scandinavian women. These groups of women may have different CVD risk factor profiles and etiologies for subfertility compared to population-based studies. To our knowledge, no longitudinal reports examine the associations of CVD risk factors with subfertility across the reproductive life span.

Our objective was to characterize the longitudinal CVD risk factor profiles of women by subfertility status in the Coronary Artery Risk Development in Young Adults (CARDIA) study, a population-based study of CVD risk factors in Black and White women. Beginning in their early twenties, CARDIA women underwent approximately four assessments of CVD risk factors. We hypothesized that women with histories of subfertility would have poorer CVD risk factor profiles over time, and that such associations would persist after adjustment for sociodemographic factors such as age, race, and education.

## Materials and methods

CARDIA is an ongoing multicenter longitudinal study conducted at four communities (Birmingham, Alabama; Chicago, Illinois; Minneapolis, Minnesota; and Oakland, California) to study CVD risk trends and clinical sequelae from young adulthood. Details of the study design, recruitment, methodology, and baseline characteristics are described elsewhere [12]. At baseline (Year 0 [Y0], 1985-1986), healthy adults (n=5,115) were recruited from the general population to be balanced on sex, race (White or Black), age (18-24 or 25-30 y) and education (high school or less, or more than high school). Data collection protocols were approved by the Institutional Review Boards of each field center with all participants providing written informed consent.

The ancillary CARDIA Women’s Study was conducted at exam year 16 (Y16) when women were approximately 40 years of age [13]. Women were asked, “Have you and a male partner ever had unprotected sexual intercourse for at least 12 months without becoming pregnant?” which is the phrasing used by the Centers for Disease Control and Prevention National Survey for Family Growth [14]. Women were also asked about reasons for subfertility. Of the 1163 women who responded, we excluded women who reported male-factor or tubal infertility (n=56) for a total of 1107 participants.

CVD risk factors were assessed at baseline and approximately every 5 years thereafter [13], for approximately four assessments between about 25 years of age to 40 years of age. Cigarette smoking was assessed by means of an interviewer-administered tobacco questionnaire and classified as current, former or never. BMI was calculated by dividing measured weight in kilograms by height in meters squared. Blood pressure was measured with participants seated and after 5 minutes of rest. The average of the second and third consecutive measurements was used for analysis. At Exam Year 15-16 (Y15-16), serum fasting glucose was measured using hexokinase coupled to glucose 6-phosphate dehydrogenase by Linco Research (St Louis, MO, USA). Lipid measurements using fasting blood samples were collected at baseline and at all follow-up examinations. Total cholesterol and triglyceride (TG) levels were measured enzymatically, high-density lipoprotein cholesterol (HDL-C) was determined after precipitation with dextran sulfate/magnesium chloride, and low-density lipoprotein cholesterol (LDL-C) was calculated by the Friedewald equation.[15]

### Statistical analysis

Participant characteristics by subfertility were defined by means (SD) or proportions as appropriate, and differences between participants in each trajectory were tested using t-tests, Wilcoxon tests, and χ^2^ analyses for continuous and categorical characteristics, respectively. To account for repeated measures of CVD risk factors between Y0 and Y16, we used generalized estimating equations with a logit link model to examine associations between subfertility (dependent variable) and CVD risk factors (independent variables). Each CVD risk factor was initially evaluated in a separate model. All models adjusted for age, race, center, and education. In a sensitivity analysis, we constructed models that adjusted for pregnancy prior to the baseline exam. We also evaluated a model with all of the risk factors combined. Analyses were performed using R version 4.4.1.

## Results

Table 1 shows the characteristics of women at baseline, when they were aged approximately 25 years of age. The majority of women had been pregnant before baseline. At Y16, approximately one-third of women (n=375) reported ever experiencing subfertility. Of the 141 women who sought medical care for subfertility, the most common etiology was irregular menstrual cycles (n=35), followed by uterine abnormalities (n=21), and endometriosis (n=18); women could report more than one etiology. At Y0, subfertile women were more likely to be black, to have had a pregnancy or live birth, to smoke, and had higher BMI and lower HDL.

**Table 1.** Characteristics of women at the baseline exam (CARDIA Exam Year 0) by any history of subfertility reported at Exam Year 16. Continuous variables compared using t-tests and categorical variables compared using χ^2^ tests.

| Table 1. Characteristics of women at the baseline exam (CARDIA Exam Year 0) by any history of subfertility reported at Exam Year 16. Continuous variables compared using t-tests and categorical variables compared using $\chi^2$ tests. | | | |
| --- | --- | --- | --- |
|  | No history of subfertility | History of subfertility | p-value |
|  | n=732 | n=375 |  |
| Age (years) | 25.3 (3.6) | 25.1 (3.7) | 0.29 |
| Black (n, %) | 351 (48%) | 238 (63.5%) | <0.0001 |
| Current cigarette use (n, %) | 164 (22.4%) | 133 (35.5%) | <0.0001 |
| Any prior pregnancy (n, %) | 372 (50.8%) | 224 (59.7%) | 0.0059 |
| Body mass index (kg/m<sup>2</sup>) | 24.6 (5.7) | 25.6 (6.1) | 0.0089 |
| Systolic blood pressure (mmHg) | 106.8 (10) | 107.3 (10.3) | 0.41 |
| Diastolic blood pressure (mmHg) | 67.3 (8.8) | 67.3 (8.9) | 0.98 |
| Fasting glucose (mg/dL) | 80.3 (12) | 81.7 (19.6) | 0.19 |
| Triglycerides (mg/dL) | 66.7 (35.8) | 68.3 (36.3) | 0.50 |
| High-density lipoprotein cholesterol (mg/dL) | 56.3 (13.1) | 53.9 (11.7) | 0.0019 |
| Low-density lipoprotein cholesterol (mg/dL) | 108.7 (29.2) | 109.4 (30.6) | 0.71 |

In models adjusted for age, race, study center, and education level, several but not all CVD risk factors were associated with subfertility. Cigarette use, higher BMI and glucose levels, and lower HDL were associated with subfertility (Table 2).

**Table 2.** Association between any history of subfertility by Exam Year 16 (dependent variable) and cardiovascular disease risk factors (independent variables) between Exam Year 0 and Exam Year 16. Each risk factor evaluated in a separate model, and all models adjust for age, race, center, and education at baseline. Odds ratios (95% confidence intervals) shown.

|  |  |  |
| --- | --- | --- |
|  | Odds ratio (95% CI) | p-value |
| Current cigarette use | 1.85 (1.50, 1.76) | <0.0001 |
| Body mass index (kg/m<sup>2</sup>) | 1.01 (1.0004, 1.02) | 0.04 |
| Systolic blood pressure (mmHg) | 1.00 (0.997, 1.01) | 0.43 |
| Diastolic blood pressure (mmHg) | 1.01 (0.999, 1.01) | 0.13 |
| Fasting glucose (mg/dL) | 1.00 (1.001, 1.01) | 0.01 |
| Triglycerides (mg/dL) | 1.001 (1.0001, 1.003) | 0.04 |
| High-density lipoprotein cholesterol (mg/dL) | 0.99 (0.986, 0.995) | <0.0001 |
| Low-density lipoprotein cholesterol (mg/dL) | 1.001 (0.999, 1.003) | 0.59 |
| C-reactive protein (mg/L) | 1.01 (0.99, 1.02) | 0.43 |

In a model with all risk factors included, smoking and HDL remained significantly associated with subfertility (Table 3); these patterns did not change with additional adjustment for any pregnancy.

**Table 3.** Association between any history of subfertility by Exam Year 16 (dependent variable) and cardiovascular disease risk factors (independent variable) between Exam Year 0 and Exam Year 16. Models adjust for age, race, center, and education at baseline **and all of the other risk factors** at Y0, Y7, Y10, and Y15. Odds ratios (95% confidence intervals) shown.

|  |  |  |
| --- | --- | --- |
|  | Odds ratio (95% CI) | p-value |
| Current cigarette use | 1.004 (1.002, 1.006) | <0.0001 |
| Body mass index (kg/m<sup>2</sup>) | 1.00 (0.99, 1.00) | 0.77 |
| Systolic blood pressure (mmHg) | 1.00 (1.00, 1.00) | 0.98 |
| Diastolic blood pressure (mmHg) | 1.00 (0.99, 1.00) | 0.23 |
| Fasting glucose (mg/dL) | 1.00 (0.99, 1.00) | 0.09 |
| Triglycerides (mg/dL) | 1.00 (1.00, 1.00) | 0.62 |
| High-density lipoprotein cholesterol (mg/dL) | 0.99 (0.995, 0.999) | 0.046 |
| Low-density lipoprotein cholesterol (mg/dL) | 1.00 (1.00, 1.00) | 0.94 |

## Discussion

In this large population-based cohort of Black and White women, subfertility was associated with cigarette use as well as higher BMI, glucose, and adverse lipid levels across the reproductive lifespan. Of these risk factors, the association between cigarette use and subfertility was the strongest. This suggests that abnormal CVD risk factor profiles during the reproductive use, particularly cigarette use, account for some of the associations previously reported between subfertility and CVD risk [1, 16-18]. Of note, continuous risk factor levels of were typically within normal range, suggesting that any impact of CVD risk factors upon subfertility may occur via profiles which are cumulative and not meeting criteria for disease. Thus, cumulative CVD risk factor levels over time may be important for CVD risk among women with histories of subfertility.

Our results are consistent with prior studies that have reported abnormalities between subfertility and CVD risk factor profiles performed at a single point in time, in selected populations. However, these studies were not consistent in which factors were associated with subfertility. In one 2020 meta-analysis, women with histories of subfertility had higher BMI, LDL, and triglycerides compared to fertile women, while glucose levels were similar in women with and without subfertility histories [4]. In contrast, in Project Viva, a cohort of pregnant women, midlife participants (∼50 years of age) with histories of subfertility (n=160) had higher glucose levels than women without histories of subfertility (n=308) [8], while other risk factors were statistically similar, and differences were not significant postmenopause. Mid-life women in the Framingham Heart Study Third Generation/Omni Cohort Exam who had histories of subfertility (n=282) had higher odds of obesity than women without histories of subfertility (n=1686) but were otherwise similar regarding their risk factors [7]. Several analyses of women participants in the National Health and Nutrition Survey (NHANES), a population-based cross-sectional study, noted that BMI, glucose, and lipids were poorer among women with histories of subfertility compared to women without these histories.[5, 6, 19]

In the present report, cigarette use was the risk factor most strongly associated with subfertility, to the extent that the significance of other CVD risk factors was markedly reduced in multivariable models including cigarette use. In one report using NHANES data, current smoking increased the odds of infertility by almost fifty-percent, even after adjustment for other sociodemographic factors [20]. Other analyses of the same participants were notable for the lack of a graded association between tobacco use and infertility [5], suggesting that even minimal exposures to cigarettes over time increased infertility risk. Exposure to cigarettes decreases ovarian reserve [21] through toxic effects upon ovarian granulosa cells [22]. Although the adverse effects of smoking for pregnancy health and CVD are established, older studies suggest that associations with infertility are not as well-recognized [23]. Dissemination of these risks may provide an additional deterrent to cigarette use in reproductive-age women.

We also found that lower levels of HDL cholesterol were associated with subfertility, even after consideration of BMI and smoking. Previous reports in NHANES have noted that higher levels of HDL are associated with lower odds of fertility [19]. Although mechanisms are speculative, HDL may serve as a substrate for key oocyte metabolites and has been linked with preimplantation embryo quality; mice with disrupted HDL metabolism have reduced fertility, which may be reversed upon restoration of cholesterol profiles [24].

Strengths of this report include the repeated measures of CVD risk factors and the population-based cohort including large numbers of Black and White women. However, this report has several limitations. Subfertility was based upon self-report, and diagnostic tests addressing the etiology of fertility were not available. Subanalysis by etiology was not possible due to sample size. The cohort was incepted approximately forty years ago, and it is possible that the CVD risk factor profiles of subfertile women differ in the present day.

We conclude that longitudinal CVD risk factor profiles are associated with subfertility, and may be a pathway through which subfertility and CVD are linked. Women should be provided with information regarding optimization of risk factors, particularly smoking, for fertility as well as future CVD risk.

## Data Availability

Data are available from the CARDIA study, dopm.uab.edu by reasonable reqeust

## Acknowledgements

Funding Statement: The Coronary Artery Risk Development in Young Adults Study (CARDIA) is conducted and supported by the National Heart, Lung, and Blood Institute (NHLBI) in collaboration with the University of Alabama at Birmingham (75N92023D00002 & 75N92023D00005), Northwestern University (75N92023D00004), University of Minnesota (75N92023D00006), and Kaiser Foundation Research Institute (75N92023D00003). This work was supported by R56HL169167 and R01HL065611. This manuscript has been reviewed by CARDIA for scientific content.

